# Cross-Cultural Adaptation and Validation of the Levenstein Perceived Stress Questionnaire (PSQ) in Ecuador

**DOI:** 10.64898/2025.12.25.25343009

**Authors:** Henry Rafael Cadena-Povea, Marco Antonio Hernández-Martínez, Amyra Gabriela Bastidas-Amador, Arlhey Didier Aguirre-Villarreal, Stalin Alexander Herrera-Falconi

**Affiliations:** Universidad Técnica del Norte, Facultad de Educación Ciencia y Tecnología, Ibarra - Ecuador

**Keywords:** Perceived Stress Questionnaire, validation, factor analysis, reliability, convergent validity, discriminant validity, factorial invariance, Ecuador, higher education, perceived stress

## Abstract

**Objective:** This study aimed to adapt and validate Levenstein’s Perceived Stress Questionnaire (PSQ) within the Ecuadorian sociocultural context. The PSQ measures perceived stress across two temporal frames: general (past two years) and specific (past month).

**Methods:** The research followed five sequential phases. *Study 1:* linguistic and cultural adaptation through standardized translation, back-translation, expert review, and a pilot test with 300 participants. *Study 2:* item refinement with an additional sample of 300 participants. *Study 3:* Exploratory Factor Analysis (EFA) with 395 participants. *Study 4:* Confirmatory Factor Analysis (CFA) with 391 participants, complemented by Structural Equation Modeling (SEM) to evaluate factorial structure, internal consistency, and convergent and discriminant validity through correlational analyses. *Study 5:* multigroup factorial invariance analysis by biological sex.

**Results:** The original questionnaire of 30 items across seven factors was refined for the Ecuadorian context into two versions: a General version with 21 items grouped into three factors, and a Specific version with 23 items distributed across four factors. Both versions demonstrated strong reliability, along with satisfactory convergent and discriminant validity.

**Conclusions:** The findings endorse the PSQ-593 as a reliable and culturally appropriate instrument for assessing perceived stress among Ecuadorian university faculty. Its validation contributes to a deeper understanding of stress as a psychological risk factor for mental health.

## I. Introduction

**(General context and relevance of stress in Ecuador)**

Ecuador is a country characterized by remarkable ethnic, cultural, and geographic diversity, reflected in a wide range of social practices, community traditions, and historical experiences that shape the daily lives of its inhabitants. This heterogeneity can significantly influence how individuals perceive, interpret, and respond to stressful events, given that the experience of stress is not solely an individual phenomenon, but also a process mediated by the sociocultural environment.

Factors such as regional differences between the Sierra, the Coast, the Amazon, and the Galápagos Islands, the contrasts between urban and rural dynamics, and the value systems specific to each ethnic and cultural group may shape the cognitive appraisal of potentially threatening or challenging situations. Stress, understood as the physiological, cognitive, and emotional response to stimuli perceived as threatening or exceeding an individual’s coping resources, is one of the most extensively studied constructs in contemporary psychology due to its close relationship with subjective well-being and physical health (Cadena et al., 2025).

From the perspective of Lazarus and Folkman (1980), perceived stress arises from a subjective assessment of environmental demands in relation to the resources available to cope with them. In this sense, stress is not simply the result of external stressors, but of how individuals interpret and evaluate those stressors. Research has shown that such interpretations differ depending on cultural, social, and personal variables, which makes it essential to study stress within specific populations. In the Ecuadorian case, this calls for a contextualized perspective that accounts for cultural factors shaping stress appraisals (Miñaca, 2023; Castilla-Gutiérrez et al., 2021; Olmos, 2013).

### Importance of Studying Stress in Health and Research

The study of stress has gained increasing relevance in psychological research and the health sciences, as it has been shown to significantly influence the development of various emotional, cognitive, and physiological disorders. Perceived stress has been linked to higher rates of anxiety and depression, as well as an elevated risk of cardiovascular disease, immune dysfunction, and metabolic problems, making it an important predictor of both quality of life and overall health (Cadena et al., 2025).

Nevertheless, there is still ongoing debate regarding the most appropriate way to conceptualize and measure this construct, since its subjective nature means that stress experiences are not evenly distributed across the population. Sociodemographic factors, such as age, gender, socioeconomic status, and cultural context, can shape how individuals assess their demands and resources, and consequently, the level of stress they report.

Within this framework, university faculty represent a particularly relevant group, as they must simultaneously cope with academic, administrative, and personal demands that can lead to high levels of strain and emotional exhaustion (Lamotta, 2023; Bolaños, 2021; Sandín, 2003). The literature indicates that the accumulation of these demands increases the likelihood of experiencing burnout, reduced performance, and difficulties in emotional regulation. This underscores the need for valid and reliable assessment tools that can identify these risks in a timely manner and guide the design of appropriate interventions.

### Particularities of Ecuadorian University Faculty

In the Ecuadorian context, university faculty face a wide range of professional demands that make them particularly vulnerable to perceived stress. These responsibilities include the planning, delivery, and assessment of courses across different academic levels; supervising students in pre-professional internships; participating in community engagement projects; and managing administrative tasks, which range from complying with institutional regulations to preparing reports and accreditation documents.

Added to these duties is the pressure to sustain a steady stream of scientific output through publications in indexed journals, participation in conferences, and securing funding for research projects—all of which intensify the cognitive and emotional load associated with academic work. Sociodemographic factors, such as sex, age, and years of professional experience, have also been shown to play a moderating role in the perception of stress. Studies highlight differences in how men and women cope with professional demands, as well as the cumulative impact of years of service on emotional exhaustion (Bolaños, 2021; Carrillo-Cruz et al., 2019; Terán, 2019; Vidal-Cisneros & Sánchez-Calva, 2018; Silva & Bonilla, 2017).

This combination of roles and demands, compounded by a context often marked by socioeconomic instability and high social expectations, contributes to heightened perceptions of strain and may negatively affect the psychological health, quality of life, and professional performance of university faculty (De Simone et al., 2021; Lee & Koh, 2020; Alvites-Huamaní, 2019).

### Cultural and Linguistic Adaptation Needs of the PSQ

The use of psychometric instruments that have not been culturally adapted or validated for the context in which they are applied can lead to misinterpretations, inaccurate assessments, and inadequate decisions in the design of intervention programs. This issue is particularly critical in countries with high sociocultural diversity, such as Ecuador, where idiomatic expressions, social norms, and the perception of stressful events can differ substantially from the contexts in which the instruments were originally developed (Guillermo & López-Regalado, 2025; López et al., 2019; Céspedes & Muñoz, 2017).

One of the most widely used instruments for measuring perceived stress is the Perceived Stress Questionnaire (PSQ), developed by Levenstein et al. (1993). This questionnaire comprises thirty items that assess daily worries, tensions, interpersonal relationships, and the sense of control over environmental demands. While the cross-cultural validity of the PSQ has been well documented in various populations, its direct application without linguistic and cultural adaptation can distort results and compromise the validity of inferences.

Therefore, it is essential to ensure semantic and conceptual equivalence through a process of translation, back-translation, and expert review, followed by rigorous statistical analyses—such as exploratory and confirmatory factor analyses—to verify the instrument’s internal structure. The guidelines of the International Test Commission (2017), along with the recommendations of Van de Vijver and Leung (2021), Hambleton et al. (2019), and Sousa and Rojjanasrirat (2010), emphasize that adaptation processes should address not only linguistic equivalence but also cultural relevance. This ensures that the scores obtained are internationally comparable and accurately reflect the experiences of the population being evaluated.

Having a psychometrically validated instrument in the Ecuadorian context represents a strategic opportunity to advance research on mental health and occupational stress in the country, as well as to support evidence-based decision-making in public policies and institutional programs. Data obtained through the use of a culturally relevant questionnaire can inform the design of psychoeducational interventions aimed at reducing stress levels among faculty, promoting healthier work environments, and preventing professional burnout. Furthermore, access to standardized and valid measures facilitates the comparison of findings across international studies, contributing to the development of a global knowledge base on the determinants of stress in higher education.

Within this framework, the primary objective of the present study was to validate the Perceived Stress Questionnaire (PSQ) in the Ecuadorian context and to adapt it linguistically and culturally to the local Spanish language. The goal was to provide a reliable and conceptually robust psychometric tool for assessing perceived stress, one that meets both research needs and the mental health challenges of the country’s population.

## II. Methodology

The study was conducted using a quantitative approach with an instrumental, cross-sectional design, aimed at examining the psychometric properties of the instrument in the Ecuadorian population. The methodology involved a systematic process of translation and cultural adaptation, followed by the administration of two pilot tests to assess and refine the clarity and relevance of the items. This was complemented by subsequent validation through exploratory factor analysis (EFA) and confirmatory factor analysis (CFA), along with a factorial invariance analysis by biological sex. The research protocol was approved by the Institutional Ethics Committee of Universidad Técnica del Norte (approval number: UTN-FECYT-CEI-2024-0000001407-C), and all procedures were conducted in accordance with the Declaration of Helsinki and local ethical guidelines from June 24 to June 25.

### Study objective

The objective of this study was to adapt and validate Levenstein’s Perceived Stress Questionnaire (PSQ) into Ecuadorian Spanish in two versions:

1. General version: designed to assess perceived stress over the past one to two years.
2. Specific version: designed to assess perceived stress over the past month.

Additionally, the study aimed to examine the psychometric properties of both versions through exploratory factor analysis (EFA), confirmatory factor analysis (CFA), and factorial invariance testing with respect to biological sex (men and women).

### Design and Approach

The present study employed an instrumental design for psychometric validation, framed within a quantitative and cross-sectional approach. The methodological process was carried out in five clearly defined and sequential phases, aiming to ensure rigor in the adaptation and validation of the Perceived Stress Questionnaire (PSQ) for the Ecuadorian context.

In the first phase, the instrument underwent linguistic and cultural adaptation to Ecuadorian Spanish, following standardized procedures of translation, back-translation, and expert review to ensure semantic and conceptual equivalence. In the second phase, a new pilot administration was conducted to make further refinements to the initial adaptation, incorporating modifications aimed at optimizing both the clarity and cultural relevance of the items.

The third phase involved conducting an Exploratory Factor Analysis (EFA) to examine the internal structure of the questionnaire and determine the most appropriate factorial configuration. In the fourth phase, a Confirmatory Factor Analysis (CFA) was performed to assess the fit of the model derived from the EFA and to verify its structural validity through various goodness-of-fit indices. Finally, in the fifth phase, a multi-group factorial invariance analysis by biological sex (men and women) was carried out to establish the instrument’s metric and structural equivalence across groups, thereby ensuring its applicability in different subpopulations.

### Participants

The study was conducted with four independent samples corresponding to the different phases of the research. In Phase 1, aimed at the initial pilot test for linguistic and cultural adaptation, 300 participants took part (41.0% men and 59.0% women), with a mean age of 33.42 years (SD = 9.85; range: 18–72 years). In Phase 2, focused on evaluating the adjustments following the adaptation, another 300 participants were included (45.7% men and 54.3% women), with a mean age of 36.18 years (SD = 10.22; range: 20–76 years).

In Phase 3, corresponding to the Exploratory Factor Analysis (EFA), the sample consisted of 395 participants (43.3% men and 56.7% women), with a mean age of 34.97 years (SD = 9.95; range: 19–75 years). Finally, in Phase 4, corresponding to the Confirmatory Factor Analysis (CFA) and the factorial invariance analysis by biological sex, 391 participants were included (43.2% men and 56.8% women), with a mean age of 34.97 years (SD = 9.96; range: 19–75 years).

Across all phases, most participants reported being either married or single, with divorced, widowed, and cohabiting individuals representing a minority. Regarding occupation, the largest group consisted of active professionals (ranging from 82.0% to 86.0% across phases), followed by students and non-practicing professionals. The most represented fields were health and education, while finance, social sciences, engineering, and service sectors had lower representation. Geographically, the majority resided in the Coastal region (70.2%–73.5% depending on the phase), followed by the Highland region (24.6%–27.8%), with a small proportion living in the Amazon region (1.8%–2.0%).

To ensure the relevance and validity of the sample, specific inclusion and exclusion criteria were established for participation in the study. Inclusion criteria required participants to be 18 years or older, to reside in Ecuador at the time of data collection, to provide voluntary consent through a signed informed consent form, and to possess sufficient reading comprehension to complete the questionnaire in Ecuadorian Spanish. Conversely, cases were excluded if the questionnaire was incomplete, contained inconsistencies, exhibited response patterns indicating a lack of engagement, or if participants did not meet the minimum age requirement or were not residing in Ecuador during the data collection period.

### Instrument: Perceived Stress Questionnaire (PSQ)

The Perceived Stress Questionnaire (PSQ), developed by Levenstein and colleagues (1993), was specifically designed for psychosomatic research as a measure of subjective perceived stress, and it is highly sensitive to temporal variations. The instrument consists of 30 items in both versions: the general version, which assesses the frequency of stress experiences over the past one to two years, and the recent or specific version, which refers to perceived stress during the past month.

Responses are registered on a 4-point Likert scale, ranging from 1 = “almost never” to 4 = “almost always.” The items include both negatively worded statements (e.g., “I feel that I have too many things to do”) and positively worded statements (e.g., “I feel well-rested”), with the latter reverse-scored for the calculation of the overall stress index. Scores are then converted into a standardized index, ranging from 0 (minimum stress level) to 1 (maximum stress level).

In its original factorial structure, the PSQ comprises seven dimensions: harassment, overload, irritability, lack of joy, fatigue, worries, and tension. These dimensions capture both cognitive and affective components of perceived stress, reflecting aspects related to external pressures, the perception of interpersonal conflicts, a sense of lack of control, and reductions in subjective well-being.

### Cultural adaptation

The adaptation of the Perceived Stress Questionnaire (PSQ) into Ecuadorian Spanish was conducted following international guidelines for the translation and cultural adaptation of psychometric instruments. First, a forward translation from English to Spanish was carried out independently by two bilingual translators with backgrounds in health sciences and applied linguistics. Subsequently, a native English speaker, blind to the original version, performed a back-translation into the source language.

Discrepancies between the original and back-translated versions were reviewed by an expert committee composed of clinical psychologists, methodology researchers, and linguistics specialists. This committee evaluated the semantic, conceptual, and cultural equivalence of the items, proposing necessary modifications to ensure their relevance within the Ecuadorian context.

The preliminary version was administered in a pilot study (Phase 1, N = 300) with the purpose of identifying potential semantic, comprehension, or cultural adequacy issues in the items. The results of this initial application highlighted the need for adjustments in terms of idiomatic expressions and clarity in the wording of certain items.

In Phase 2 (N = 300), the revised version of the questionnaire was administered, allowing for the evaluation of the effectiveness of the linguistic and cultural modifications before proceeding to the factorial analyses. This process ensured that the Ecuadorian version of the PSQ remained faithful to the original construct, while being comprehensible and culturally appropriate for the target population.

### Statistical Procedure

The statistical analysis was conducted in several sequential stages. First, assumptions regarding internal consistency and item adequacy were verified. To do this, Cronbach’s alpha was calculated for both versions of the questionnaire. In the general version, only item 25 showed a low item–total correlation; however, it was retained for subsequent factorial analyses. In the specific version, all items reached satisfactory values, with no problematic elements identified. Additionally, multicollinearity among items was examined, and no evidence of redundancy was found that could compromise the instrument’s validity.

Subsequently, the factorability of the data matrix was assessed using the Kaiser-Meyer-Olkin (KMO) index, which yielded a value above 0.70, and Bartlett’s test of sphericity, which was statistically significant (p < .001). These results confirmed the suitability of the data for factorial analysis.

In the third phase, an exploratory factor analysis (EFA) was conducted using the maximum likelihood method. Factor extraction employed oblique rotation (Oblimin), given the assumed intercorrelation among the dimensions of perceived stress. Retention criteria were established as factor loadings equal to or greater than 0.25, consistent with the threshold reported in the original test, as well as the absence of problematic cross-loadings that could compromise the factorial clarity of the items.

In the fourth phase, a confirmatory factor analysis (CFA) was undertaken to evaluate the structural validity of the models identified through the EFA. The goodness of fit was determined using the chi-square statistic (χ²) along with several complementary indices: Comparative Fit Index (CFI), Tucker–Lewis Index (TLI), Root Mean Square Error of Approximation (RMSEA), and Standardized Root Mean Square Residual (SRMR).

Thereafter, factorial invariance analysis was conducted with respect to biological sex (men and women). This procedure involved the progressive estimation of configural, metric, scalar, and strict models. Invariance was evaluated by comparing model fit indices, with changes of ΔCFI ≤ 0.01 and ΔRMSEA ≤ 0.015 considered acceptable.

Finally, the internal reliability of the scales was assessed using two complementary coefficients: Cronbach’s alpha, as the traditional measure of internal reliability, and McDonald’s omega, a more robust estimator that does not rely on the assumption of tau-equivalence. This dual approach allowed for a more accurate assessment of the internal reliability of both versions of the questionnaire.

## III. Results

### General Version– Exploratory Factor Analysis (AFE)

The exploratory factor analysis (EFA) of the general version of the PSQ, administered with reference to the past year or two, revealed adequate indicators of the factorability of the correlation matrix. The Kaiser-Meyer-Olkin (KMO) reached 0.925, reflecting excellent sample adequacy for factor analysis. Additionally, Bartlett’s test of sphericity was statistically significant (χ² = 2932.394, df = 210, p < 0.001), confirming that the data matrix was suitable for factor extraction. This significance is consistent with the sample size (N = 395), as Bartlett’s test tends to yield high χ² values when the number of participants is substantial, thereby increasing the statistical power of the analysis.

Factor extraction was performed using principal axis factoring with oblique (Oblimin) rotation and Kaiser normalization, converging after seven iterations. This process made it possible to identify a three-factor structure with eigenvalues above 1, which together accounted for 49.776% of the total variance: the first factor explained 33.943%, the second 10.764%, and the third 5.070%.

Regarding the item distribution, Factor 1 was the most extensive, encompassing the majority of items. This factor appeared to be composed of two subfactors: the first subfactor included items G2, G4, G11, G16, G19, G28, and G30, while the second subfactor consisted of items G8, G15, G18, G20, and G26. Factor 2 grouped items G7, G10, G13, G17, and G21, and Factor 3 comprised items G5, G6, G12, and G23. Overall, a four-factor model is observed.

The extracted communalities showed adequate values for most items, ranging from 0.23 to 0.67, indicating that the retained factors account for a substantial proportion of the variance of each question. Moreover, the internal reliability of the scale was high, with a Cronbach’s α of 0.894 for the total of the 21 retained items, supporting the reliability of the identified factorial structure.

### General Version – Confirmatory Factor Analysis (CFA)

The confirmatory factor analysis (CFA) was conducted to test the three-factor structure identified in the EFA. The overall fit indices indicated an adequate performance of the model. The chi-square statistic was significant (χ² = 403.438, df = 183, p < .001), with a χ²/df ratio of 2.205—a value considered acceptable and within the recommended range for models with large samples.

Regarding comparative fit indices, the model yielded CFI = 0.920, IFI = 0.921, TLI = 0.908, and NFI = 0.864, all of which exceed or approach the 0.90 threshold typically suggested to indicate good fit. The RMSEA = 0.056 (90% CI: 0.048–0.063) also fell within the expected range, with a PCLOSE value of 0.103, suggesting that the hypothesis of close fit cannot be rejected. Finally, the SRMR = 0.056, which complements absolute error indices, further supports the adequacy of the model.

It is important to note that the model was validated through structural equation modeling (SEM), given that one of the three factors identified in the EFA exhibited two internal subfactors. This methodological decision made it possible to evaluate the validity of the four-factor structure more accurately, taking into account both the overall relationships among factors and the internal differentiation of the first factor.

The four-factor structure, with the first factor subdivided into two subdimensions, fits the data satisfactorily, supporting the structural validity of the general version of the PSQ in the Ecuadorian population.

### General Version – Factorial Invariance

The multigroup factorial invariance analysis showed that the four-factor configurational model had a good fit, confirming that the factorial structure is consistent across the groups evaluated. When constraints were imposed on the factor loadings (metric invariance), the fit remained stable (ΔCFI = 0.002), supporting the equivalence of the loadings across groups.

Subsequently, scalar invariance—which adds constraints on intercepts—demonstrated a satisfactory fit without significant deterioration (ΔCFI < 0.01; ΔRMSEA < 0.015), thus allowing for the comparison of latent means across groups. When testing for strict invariance, which equalizes measurement residuals, a slight deterioration in fit was observed (ΔCFI = 0.011), marginally above the conservative criterion proposed by Cheung and Rensvold (2002). However, the ΔRMSEA remained below 0.015, suggesting that the model can be considered practically invariant in its measurement errors.

Overall, the findings support the validity of cross-group comparisons for the general version of the PSQ in the Ecuadorian context.

### Specific Version – Exploratory Factor Analysis (EFA)

The exploratory factor analysis was conducted on a sample of 395 participants. Sampling adequacy was excellent, with a KMO index of 0.928, described as “marvelous” by Kaiser (1974), confirming the suitability of the data for factor analysis. Bartlett’s test of sphericity was statistically significant (χ²(253) = 3596.27, p < .001), rejecting the null hypothesis of an identity matrix and supporting the factorability of the correlation matrix.

The extracted communalities ranged from 0.285 to 0.692, values that are considered low for some items but deemed acceptable for retention according to the ≥ 0.25 criterion used in the original PSQ version. Despite these moderate communalities, the factor loadings were highly satisfactory, leading to the decision to retain the items due to their significant contribution to the construct measured by the instrument. Additionally, the internal consistency of the scale was high, with a Cronbach’s alpha of 0.911 for the 23 items, confirming excellent reliability for the specific version of the instrument.

These results provide strong evidence to proceed with factor extraction and support the construct validity of the specific version of the PSQ-593. The factor extraction, conducted using principal axis factoring with oblique rotation (Oblimin) and Kaiser normalization, converged in 13 iterations and revealed a structure composed of four factors with eigenvalues greater than 1, which together explained 54.970% of the total variance. The first factor accounted for 34.893% of the variance, the second for 10.337%, the third for 5.293%, and the fourth for 4.447%.

The EFA revealed a four-factor solution with adequate internal consistency and conceptual coherence. The first factor consisted of items E2, E3, E4, E8, E11, E14, E16, E18, and E26; the second factor included items E1, E7, E10, E13, E17, E21, and E25; the third factor encompassed items E5, E6, E20, E23, and E24; and the fourth factor comprised items E9 and E22. These results demonstrate a coherent and statistically robust factorial structure for the specific version of the PSQ-593.

### Specific Version – Confirmatory Factor Analysis

The confirmatory factor analysis (CFA) was conducted on a sample of 391 participants, with the aim of evaluating the adequacy of the four-factor model obtained in the EFA. The global fit indices indicated a satisfactory model fit: χ²(222) = 479.930, p < .001, with a χ²/df ratio of 2.162—a value considered optimal for models of this level of complexity. Comparative fit indices showed an NFI = 0.868 (slightly below the recommended threshold of 0.90), whereas IFI = 0.925, TLI = 0.913, and CFI = 0.924 exceeded the acceptability criterion, demonstrating that the model adequately reproduces the observed covariance matrix. The approximation error was low (RMSEA = 0.055, 90% CI [0.048, 0.061], PCLOSE = 0.127), falling within the range of excellence (< 0.06).

The χ²/df ratio close to 2, together with the low RMSEA, reflects a parsimonious and well-specified model. This ensures an appropriate balance between fit and complexity, while avoiding overparameterization. Although the NFI fell slightly below the 0.90 threshold, this index is sensitive to sample size and the number of estimated parameters. Therefore, marginally lower values do not necessarily indicate poor fit.

Taken together, the results confirm that the four-factor solution is both statistically robust and theoretically consistent, providing strong support for the construct validity of the specific version of the PSQ in the Ecuadorian context.

### Specific Version – Factorial Invariance

In order to evaluate the equivalence of the factorial structure of the specific version of the PSQ across the analyzed groups, a multigroup factorial invariance analysis was conducted, following a hierarchical approach. The configural model showed satisfactory fit, confirming that the four-factor structure is consistent across the evaluated groups.

When imposing the restriction of factor loadings (metric invariance), the fit remained stable (ΔCFI = 0.002), indicating that the items contribute equivalently to the measurement of the construct in the different groups. Scalar invariance, which adds the equality of intercepts, showed no variation in fit (ΔCFI = 0.000). This robustly confirms intercept equivalence, validating the comparison of latent means.

However, when evaluating strict invariance, which involves equality of measurement residuals, a considerable deterioration in fit was observed (ΔCFI = −0.018), along with an increase in RMSEA of 0.02. This indicates that strict invariance was not achieved.

Overall, the results support configural, metric, and scalar invariance, allowing for comparisons of latent means across groups. Nevertheless, the lack of strict invariance suggests that measurement errors are not fully equivalent across groups, and therefore, comparisons of variances and covariances should be interpreted with caution.

### Reliability

The internal consistency of the adapted versions of the PSQ-593 was satisfactory. For the general version, the overall Cronbach’s alpha was 0.894, and the omega coefficient was 0.890—indicating excellent global reliability. At the factor level, alpha coefficients ranged from 0.718 (External demands) to 0.819 (Interpersonal distress), with very similar omega values. This confirms the stability of the reliability estimates.

In the specific version, the overall reliability was slightly higher (α = 0.911; ω = 0.909), supporting even more the robustness of the instrument for assessing perceived stress over recent periods. Factors presented consistent alpha and omega coefficients: External Demands (α = 0.798; ω = 0.799), Well-being (α = 0.790; ω = 0.791), and Interpersonal Distress (α = 0.882; ω = 0.882)—all within ranges indicating high reliability.

For the Emotional Distress factor, which consists of only two items, neither Cronbach’s alpha nor omega coefficients were reported. Instead, the Spearman correlation between the two items was calculated, yielding a moderate value (ρ = 0.476). Although this result falls below conventional alpha thresholds, it aligns with the literature, which notes that Cronbach’s alpha tends to underestimate reliability in two-item scales. Additionally, item-to-item correlation is a more appropriate indicator in such cases. Moreover, the observed correlation is sufficiently high to rule out extreme item independence, confirming the absence of multicollinearity and thereby preserving the factor’s unique contribution to the measured construct.

The results indicate that the adapted versions of the PSQ demonstrate excellent internal consistency and serve as psychometrically reliable tools for assessing perceived stress in the Ecuadorian population. Notably, the specific version showed slightly higher overall and factor-level reliability indices, suggesting that it may be a more sensitive and precise measure for capturing perceived stress over recent periods.

### Construct Validity

The exploratory and confirmatory factor analyses conducted on the Ecuadorian sample confirmed the suitability of a four-factor structure for both the general version (perceived stress over 1–2 years) and the specific version (recent stress, past month). The identified factors—External Demands, Well-being, Interpersonal Distress, and Emotional Distress—show strong factor loadings that are consistent with the theoretical model of the PSQ proposed by Levenstein.

In the specific version, the factors appeared more clearly defined and conceptually homogeneous. External Demands exclusively grouped items related to objective overload, time pressure, and social demands, whereas Emotional Distress encompassed somatic, cognitive, and affective indicators of stress. In contrast, in the general version, some items reflecting emotional responses (e.g., fatigue, irritability) clustered together with External Demands, suggesting a greater integration between perceived load and emotional reaction when stress is assessed over longer periods.

These findings support the construct validity in both versions, confirming that the instrument consistently measures the central dimensions of perceived stress within the Ecuadorian context.

## Discussion

### Contextualization and International Comparison

The validation of the PSQ-593 confirms the relevance of the instrument as a sensitive and multidimensional measure of perceived stress, while also highlighting the importance of considering sociocultural factors in its adaptation process. The results align with findings reported in other linguistic and cultural contexts. For instance, Sanz-Carrillo et al. (2002), in the Spanish adaptation, found a factorial structure consistent with the original version, making editorial adjustments to ensure semantic equivalence. Meanwhile, Fliege et al. (2005), in the German validation, observed variations in the loadings of items related to perceived control, suggesting a cultural component in the experience of stress. Similarly, Escobar-Córdoba et al. (2011) emphasized, in the Colombian population, the need to account for social and linguistic context, and Leung et al. (2010) reported, in the Chinese adaptation, the importance of adjusting items linked to social interaction in accordance with collectivist values.

### Factorial Structure and Invariance

In the general version as well as in the specific version of the PSQ-593, the four-factor structure identified through exploratory factor analysis was confirmed by confirmatory factor analysis, with satisfactory overall fit indices in both models (χ²/df < 3; CFI > 0.90; RMSEA < 0.06). These findings provide strong support for the construct validity of both versions and demonstrate that the core conceptual structure of the instrument remains stable, regardless of the reference period used to measure perceived stress.

Furthermore, the multigroup factorial invariance analysis conducted on the specific version showed configural, metric, and scalar equivalence, indicating that the factorial organization, item loadings, and intercepts are consistent across the groups analyzed. This allows for valid comparisons of latent means. Although strict invariance was not achieved (ΔCFI = −0.018; ΔRMSEA = +0.02), this limitation is common in psychometric instruments and does not invalidate the use of the PSQ-593 for mean comparisons, though caution is advised when interpreting variances, covariances, and measurement errors.

Overall, these results confirm that both versions of the PSQ-593 are psychometrically robust and adequately capture the experience of stress in the Ecuadorian population, while also supporting their use in both retrospective research (general version) and studies requiring assessment of recent stress (specific version).

### Item Grouping and Structural Stability

One of the most notable aspects of this study is that, although both the general and specific versions of the PSQ-593 retained a four-factor structure, the grouping of items showed slight variations that reflect the instrument’s sensitivity to the evaluated time frame. In the External demands factor, for example, both versions included items that reflect overload and pressure (E2, E4, E11, E16), but the general version also incorporated indicators of overall tension and fatigue (E3, E8, E14, E18, E26), while the specific version grouped more situational items related to external pressure and deadlines (E19, E28, E30). Likewise, within the Well-being factor, the general version integrated items related to rest and relaxation (E1, E25), which do not appear in the specific version, probably because in short-term assessments the perception of immediate affective states (E7, E10, E13, E17, E21) tends to prevail. In interpersonal Distress, the specific version included the frustration item (E12) and excluded the social criticism item (E24), suggesting that recent personal experiences based on conflicts carry more weight than the perception of being judged by others. In turn, the emotional Distress factor expanded from two items in the general version (E9, E22) to five in the specific version (E8, E15, E18, E20, E26), showing that recent stress manifests through a wider range of emotional symptoms. These variations confirm that, even though the multidimensional structure remains stable, the distribution of items adjusts to the reference time frame, providing a more contextualized and sensitive measurement of stress experiences in each version.

### Reliability and International Comparison

The adapted versions of the PSQ-593 demonstrated excellent internal consistency. The general version showed a Cronbach’s alpha of 0.894 and an omega coefficient of 0.890, while the specific version yielded even higher values (α = 0.911; ω = 0.909), indicating a highly precise and stable measurement. The individual factors also showed satisfactory reliability levels, all of them above the 0.70 threshold recommended by Nunnally and Bernstein (1994). In the case of Emotional Distress, which was composed of two items, the Spearman correlation (ρ = 0.476) was sufficient to justify its inclusion, in consonance with the original proposal by Levenstein et al. (1993), which also considered a two-items dimensions. These results surpass the alpha coefficients reported in the original validation of the PSQ (0.80–0.85) and are consistent with later adaptations in other countries, where values are usually found within the 0.80 to 0.90 range (Fliege et al., 2005; Sanz-Carrillo et al., 2002; Moretti & Medrano, 2014). This finding confirms confirms that the Ecuadorian adaptation not only preserves the reliability of the instrument but in some cases enhances it, providing a robust tool for assessing perceived stress across different research and clinical practice settings.

### Theoretical and Sociocultural Implications

The findings of this research can be interpreted in light of Lee’s (2010) concept of the “superstress syndrome,” which affirms that in contemporary society, the constant increase in work, social, and family demands turns stress into a chronic phenomenon, with significant consequences for psychological and physical well-being. In the Ecuadorian context, this situation is intensified by the multiple roles assumed by professionals, the socioeconomic inestability, and high social expectations, creating a pattern of perceived stress that extends beyond the workplace and affects personal and community life. The fact that the adaptation of the PSQ required linguistic and semantic adjustments to accurately reflect these experiences supports the hypothesis that the experience of stress is influenced by sociocultural factors, and that its measurement must incorporate these particularities to ensure validity.

The results of this validation carry significant implications for both research and professional practice. The confirmation of construct validity and the high reliability of both versions of the PSQ-593 support their use in epidemiological studies, psychoeducational interventions, and stress prevention programs across different settings. The specific version, when showing slightly higher reliability indices, is particularly useful for monitoring short-term interventions and for assessing the impact of recent events on perceived stress. The absence of strict invariance suggests that future research should review items with greater residual variability across groups to optimize precision and enhance full comparability of scores. This study constitutes a pioneering contribution in the Ecuadorian context, as no locally validated instruments were previously available to measure perceived stress. Consequently, the PSQ-593 not only provides a robust and culturally relevant tool but also paves the way for the validation of other instruments related to stress, anxiety, coping, and resilience, contributing to the development of a country-specific theoretical and empirical body of knowledge.

### Strengths, Limitations, and Prospective Use

One strength of this study is in its rigorous methodology, which included both exploratory and confirmatory factor analyses, as well as an evaluation of factorial invariance, supporting the robustness of the findings. The amplitude and representativeness of the sample further reinforce the generalizability of the results. As a limitation, the absence of previously validated instruments in the Ecuadorian context that would allow for assessing convergent or discriminant validity is acknowledged, although this highlights the pioneering nature of this work. The findings position the PSQ-593 as a valuable resource for future research, psychoeducational intervention programs, and the development of public health policies aimed at early detection and management of stress in work, educational, and community settings.

**Figure 1.**
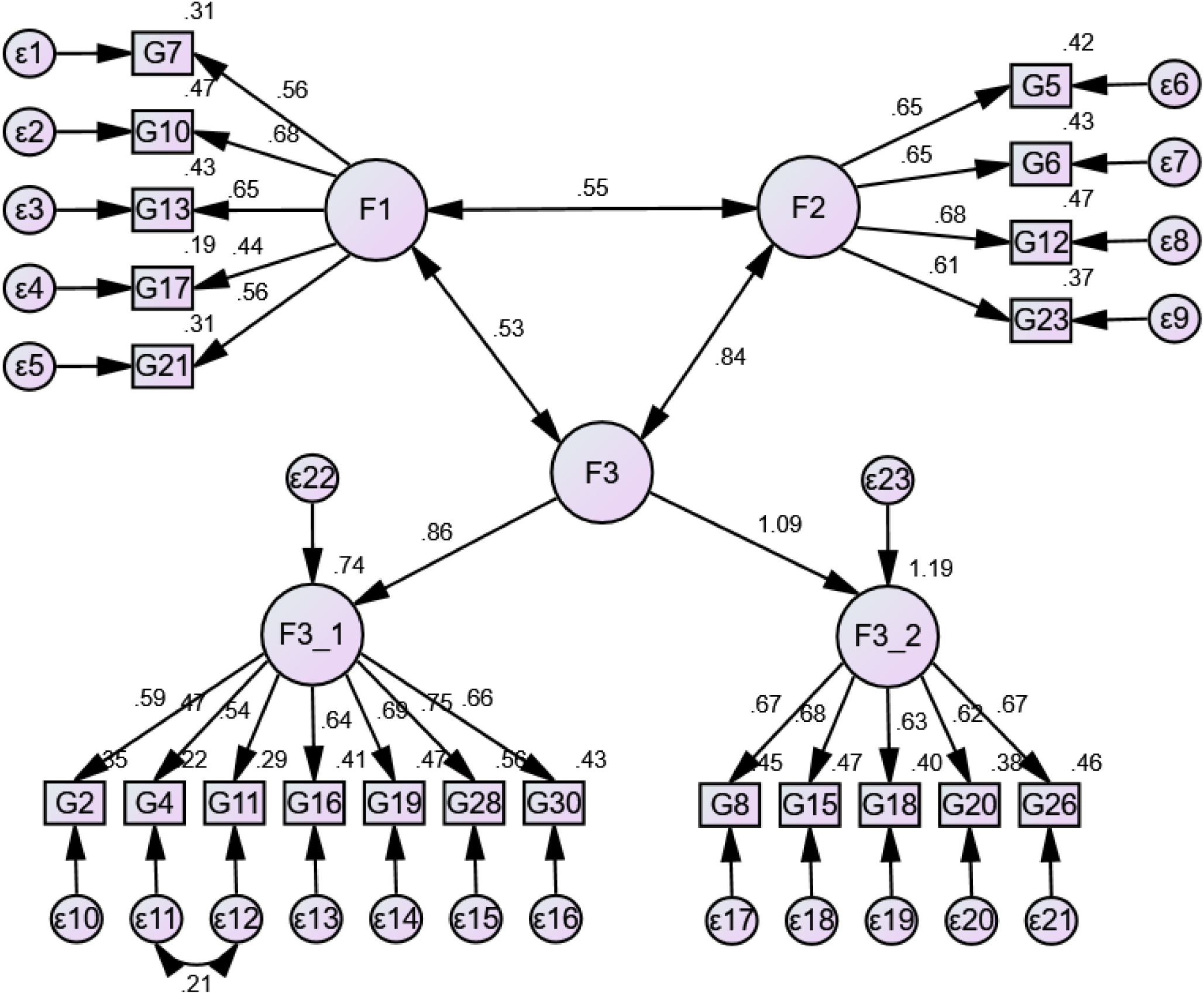
Confirmatory factor model of the general version with 4 factors.

**Figure 2.**
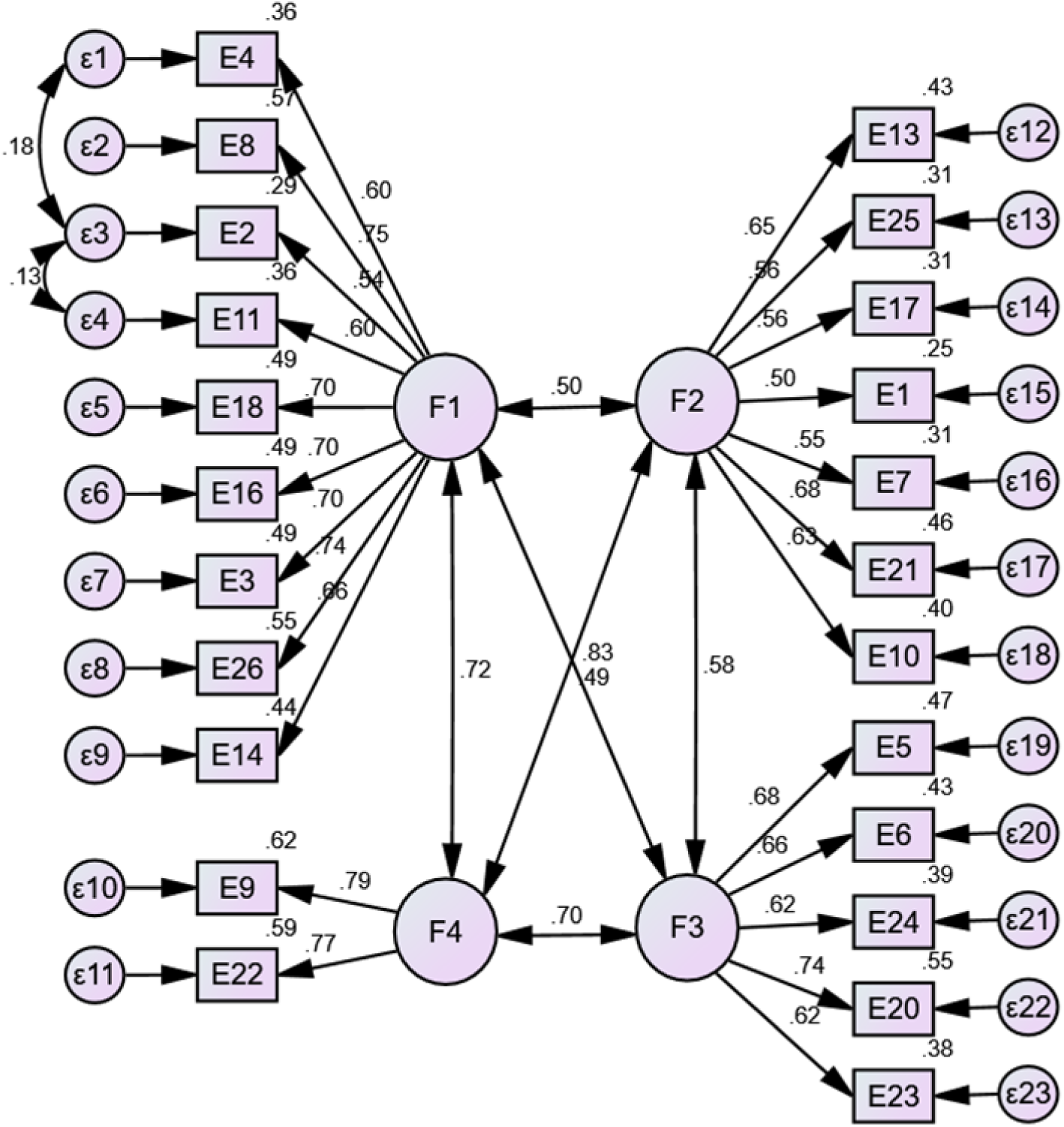
Confirmatory Factor Model of the Specific Version.

**Table 1.**
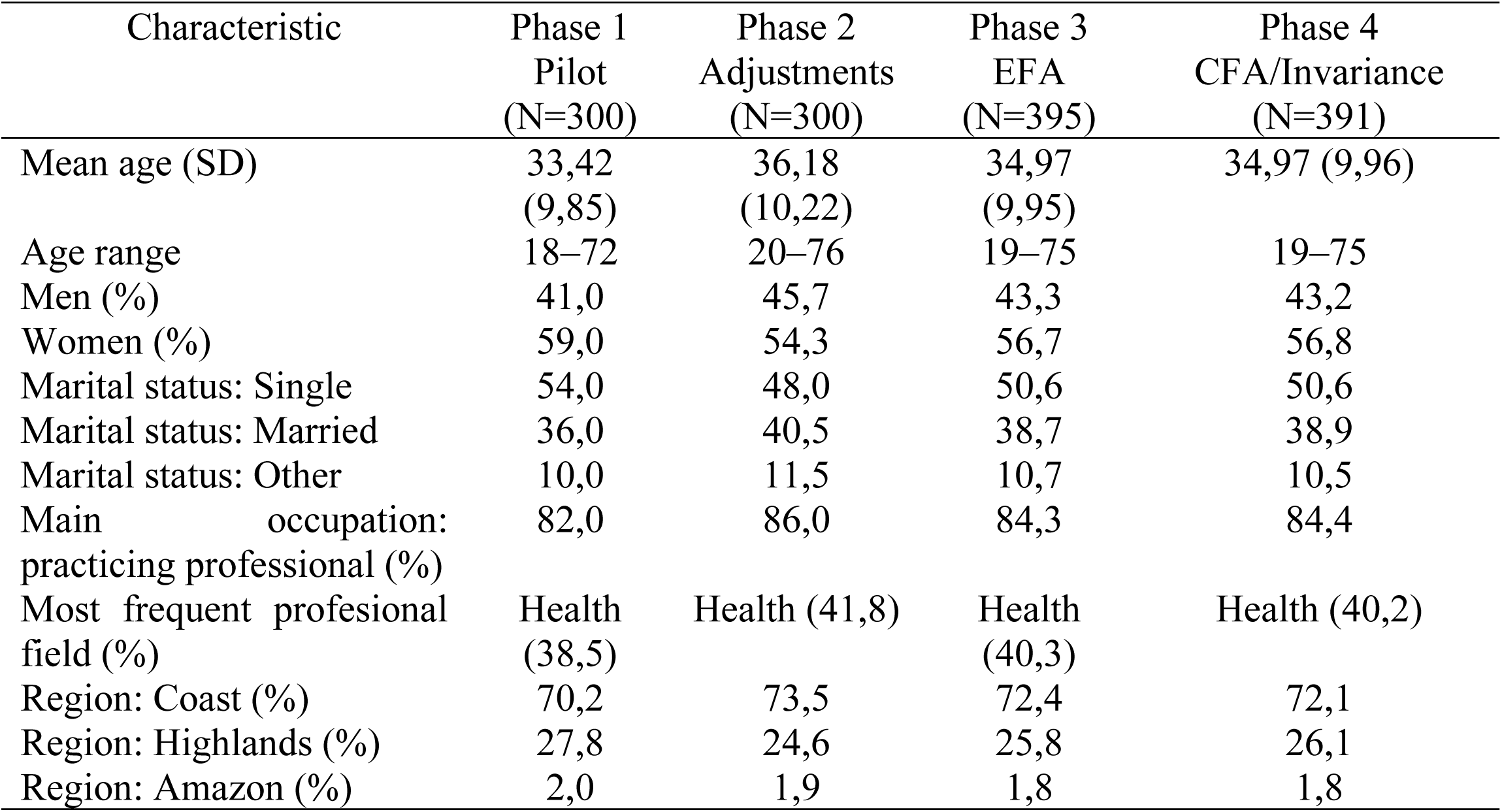
Sociodemographic Characteristics of Participants by Phase and PSQ.

| Characteristic | Phase 1<br>Pilot<br>(N=300) | Phase 2<br>Adjustments<br>(N=300) | Phase 3<br>EFA<br>(N=395) | Phase 4<br>CFA/Invariance<br>(N=391) |
| --- | --- | --- | --- | --- |
| Mean age (SD) | 33,42<br>(9,85) | 36,18<br>(10,22) | 34,97<br>(9,95) | 34,97 (9,96) |
| Age range | 18–72 | 20–76 | 19–75 | 19–75 |
| Men (%) | 41,0 | 45,7 | 43,3 | 43,2 |
| Women (%) | 59,0 | 54,3 | 56,7 | 56,8 |
| Marital status: Single | 54,0 | 48,0 | 50,6 | 50,6 |
| Marital status: Married | 36,0 | 40,5 | 38,7 | 38,9 |
| Marital status: Other | 10,0 | 11,5 | 10,7 | 10,5 |
| Main occupation:<br>practicing professional (%) | 82,0 | 86,0 | 84,3 | 84,4 |
| Most frequent profesional<br>field (%) | Health<br>(38,5) | Health (41,8) | Health<br>(40,3) | Health (40,2) |
| Region: Coast (%) | 70,2 | 73,5 | 72,4 | 72,1 |
| Region: Highlands (%) | 27,8 | 24,6 | 25,8 | 26,1 |
| Region: Amazon (%) | 2,0 | 1,9 | 1,8 | 1,8 |

**Table 2.**
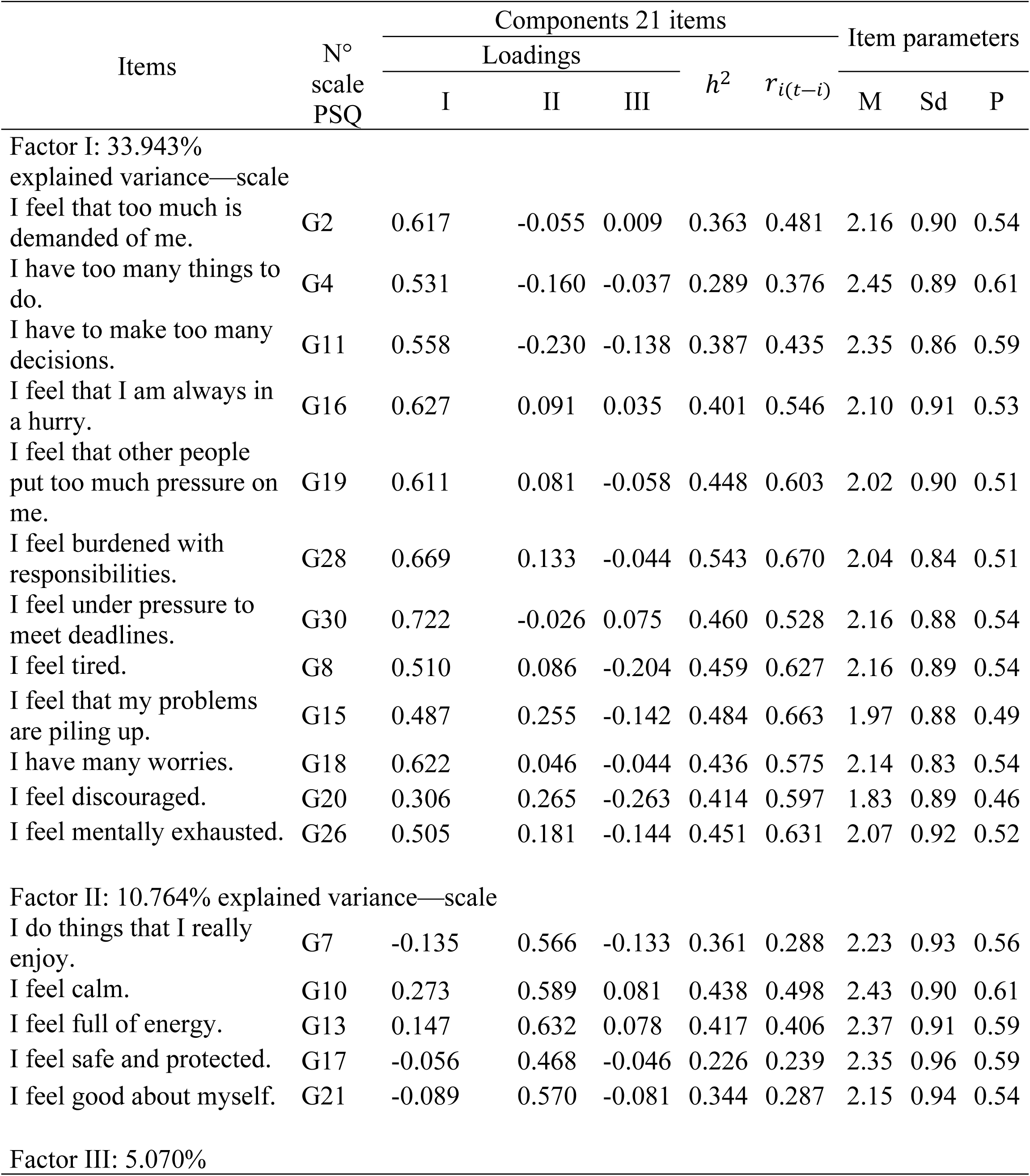
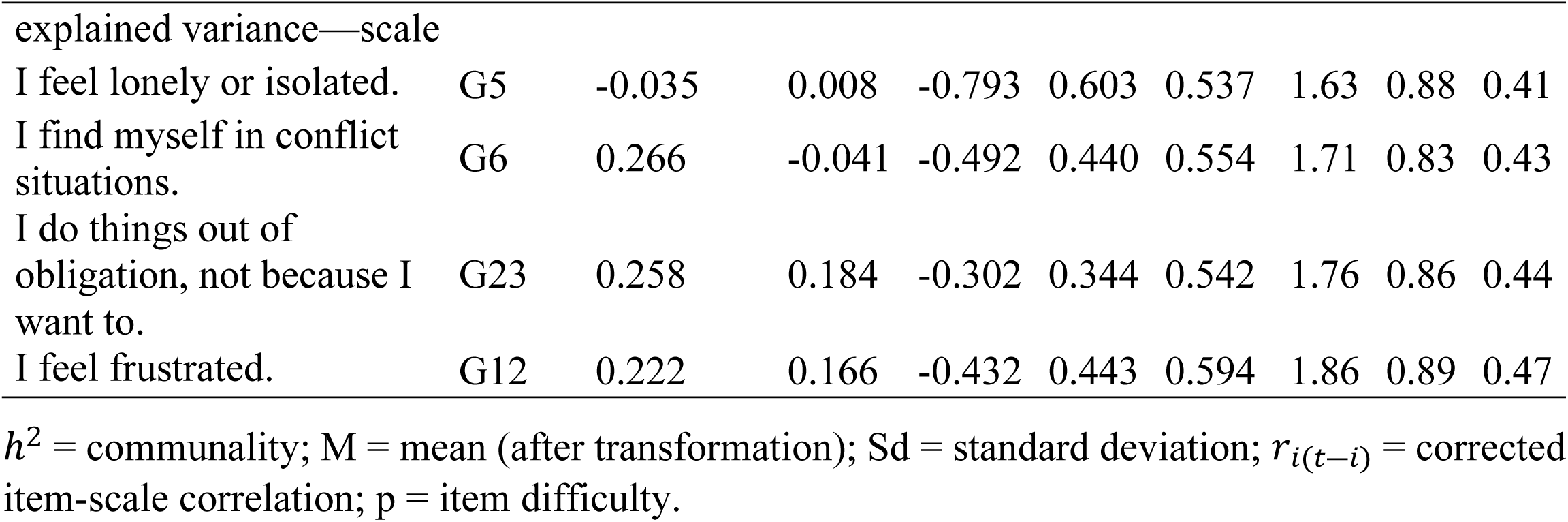
Exploratory factor analysis with Oblimin-rotation of the 21 PSQ-593 items.

**Table 3.**
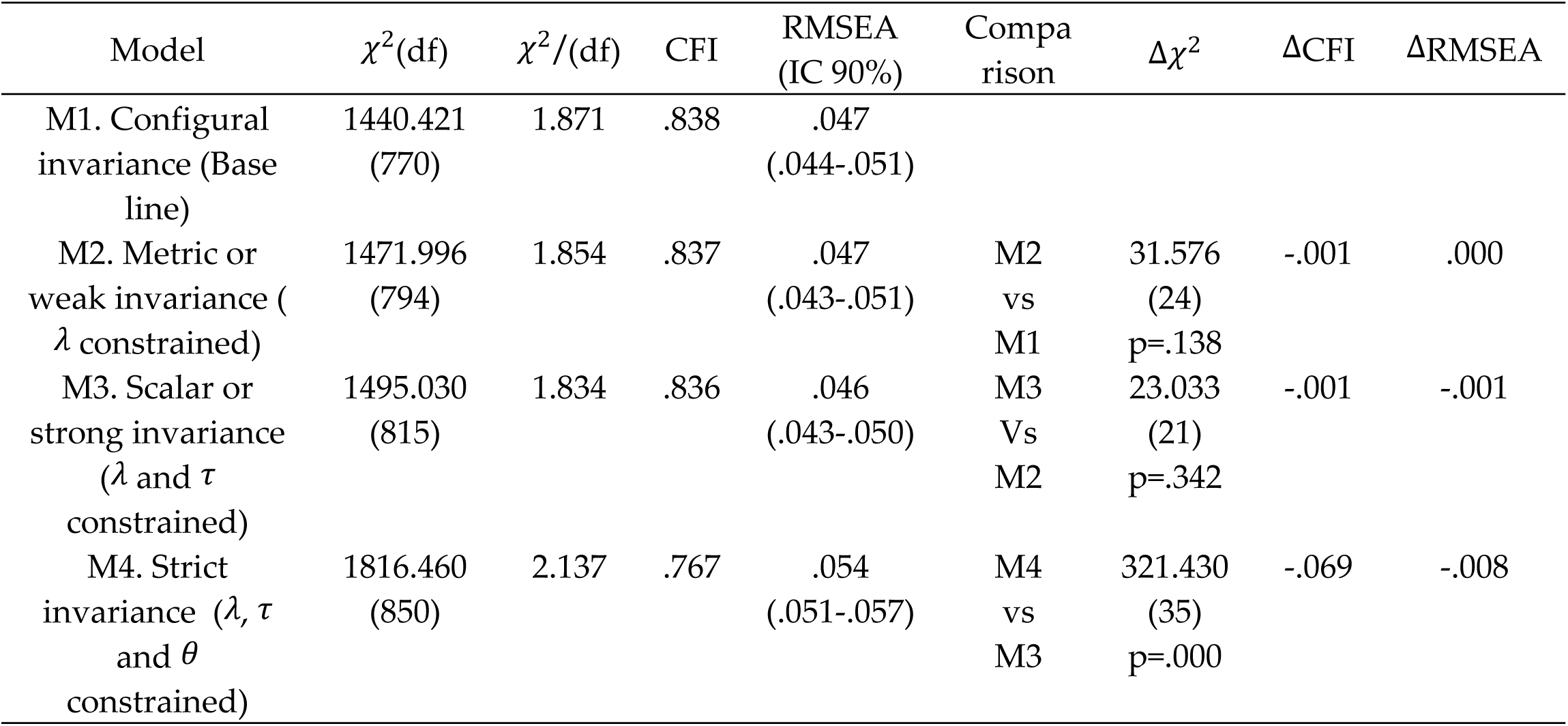
Fit indices and changes for the invariance analysis by sex (general version).

**Table 4.** Exploratory factor analysis with Oblimin-rotation of the 23 PSQ-593 items.

| Items | N°<br>scale<br>PSQ | Components 23 items |  |  |  |  |  | Item parameters |  |  |
| --- | --- | --- | --- | --- | --- | --- | --- | --- | --- | --- |
| | | Loadings | | | | $h^2$ | $r_{i(t-i)}$ | M | Sd | p |
|  |  | I | II | III | IV |  |  |  |  |  |
| Factor I: 34.893 %<br>explained variance—scale |  |  |  |  |  |  |  |  |  |  |
| I have felt that too much is demanded of me. | E2 | 0.581 | -0.035 | -0.112 | 0.031 | 0.382 | 0.458 | 2.15 | 0.93 | 0.54 |
| I have felt irritable or in a bad mood. | E3 | 0.471 | 0.162 | -0.284 | -0.002 | 0.523 | 0.662 | 1.95 | 0.92 | 0.49 |
| I have had too many things to do. | E4 | 0.726 | -0.035 | 0.042 | -0.020 | 0.506 | 0.475 | 2.36 | 0.95 | 0.59 |
| I have felt tired. | E8 | 0.583 | 0.193 | -0.106 | -0.070 | 0.571 | 0.675 | 2.21 | 0.90 | 0.55 |
| I have had to make too many decisions. | E11 | 0.536 | -0.002 | 0.023 | -0.213 | 0.422 | 0.523 | 2.29 | 0.88 | 0.57 |
| I have felt tense. | E14 | 0.448 | 0.154 | -0.137 | -0.109 | 0.432 | 0.607 | 2.02 | 0.90 | 0.50 |
| I have felt that I am always in a hurry | E16 | 0.480 | -0.035 | -0.184 | -0.190 | 0.483 | 0.594 | 2.16 | 0.95 | 0.54 |
| I have had many worries. | E18 | 0.502 | 0.088 | -0.096 | -0.180 | 0.484 | 0.624 | 2.21 | 0.91 | 0.55 |
| I have felt mentally exhausted. | E26 | 0.454 | 0.157 | -0.137 | -0.199 | 0.530 | 0.676 | 2.08 | 0.96 | 0.52 |
| Factor II: 10.337 %<br>explained variance—scale |  |  |  |  |  |  |  |  |  |  |
| I have felt rested. | E1 | 0.162 | 0.523 | 0.045 | 0.059 | 0.284 | 0.377 | 2.77 | 0.86 | 0.69 |
| I have done things that I really enjoy. | E7 | -0.203 | 0.518 | -0.082 | -0.113 | 0.339 | 0.318 | 2.32 | 0.91 | 0.58 |
| I have felt calm. | E10 | 0.159 | 0.508 | -0.102 | -0.026 | 0.404 | 0.541 | 2.44 | 0.87 | 0.61 |
| I have had a lot of energy. | E13 | -0.023 | 0.646 | 0.009 | -0.078 | 0.452 | 0.437 | 2.38 | 0.92 | 0.59 |
| I have felt safe and protected. | E17 | -0.029 | 0.541 | -0.073 | 0.026 | 0.310 | 0.363 | 2.31 | 0.92 | 0.58 |
| I have felt good about myself. | E21 | -0.242 | 0.517 | -0.382 | -0.046 | 0.524 | 0.458 | 2.10 | 0.93 | 0.52 |
| I have felt relaxed and worry-free. | E25 | 0.081 | 0.624 | 0.143 | -0.020 | 0.364 | 0.361 | 2.78 | 0.88 | 0.69 |
| Factor III: 5.293 %<br>explained variance—scale |  |  |  |  |  |  |  |  |  |  |
| I have felt lonely or isolated. | E5 | -0.013 | -0.037 | -0.630 | -0.205 | 0.536 | 0.570 | 1.62 | 0.89 | 0.41 |
| I have found myself in conflicting situations. | E6 | 0.303 | -0.065 | -0.574 | 0.056 | 0.505 | 0.561 | 1.72 | 0.86 | 0.43 |
| I have felt discouraged. | E20 | 0.174 | 0.087 | -0.458 | -0.169 | 0.504 | 0.641 | 1.80 | 0.87 | 0.45 |
| I have done things out of obligation, not because I wanted to. | E23 | 0.085 | 0.170 | -0.381 | -0.142 | 0.375 | 0.558 | 1.84 | 0.87 | 0.46 |
| I have felt criticized or judged. | E24 | 0.156 | 0.056 | -0.523 | -0.008 | 0.409 | 0.544 | 1.78 | 0.85 | 0.45 |
| | | Loadings | | | | $h^2$ | $r_{i(t-i)}$ | M | Sd | p |
|  |  | I | II | III | IV |  |  |  |  |  |
| Factor IV: 4.447 %<br>explained variance—scale |  |  |  |  |  |  |  |  |  |  |
| I have feared not to<br>achieve my goals. | E9 | 0.019 | -0.016 | -0.015 | -0.822 | 0.692 | 0.582 | 2.08 | 0.96 | 0.52 |
| I have been afraid of<br>the future. | E22 | 0.134 | 0.031 | -0.005 | -0.635 | 0.522 | 0.564 | 2.03 | 0.98 | 0.51 |

$h^2$ = communality; M = mean (after transformation); Sd = standard deviation; $r_{i(t-i)}$ = corrected item-scale correlation; p = item difficulty.

**Table 5.** Fit Indices and Changes for the Invariance Analysis by Sex (specific version).

| Model | $\chi^2(\text{df})$ | $\chi^2/(\text{df})$ | CFI | RMSEA<br>(IC 90%) | Comparison | $\Delta\chi^2$ | $\Delta\text{CFI}$ | $\Delta\text{RMSEA}$ |
| --- | --- | --- | --- | --- | --- | --- | --- | --- |
| M1. Configural invariance (Base line)) | 856.095<br>(444) | 1.928 | .884 | .049<br>(.044-.054) |  |  |  |  |
| M2. Metric or weak invariance ($\lambda$ constrained)) | 883.860<br>(463) | 1.909 | .882 | .048<br>(.043-.053) | M2<br>vs<br>M1 | 27.765<br>(19)<br>p=.088 | -.002 | -.001 |
| M3. Scalar or strong invariance ($\lambda$ and $\tau$ constrained) | 893.501<br>(473) | 1.889 | .882 | .048<br>(.043-.053) | M3<br>Vs<br>M2 | 9.640<br>(10)<br>p=.473 | -.000 | -.000 |
| M4. Strict invariance ($\lambda$, $\tau$, and $\theta$ constrained) | 981.455<br>(498) | 1.971 | .864 | .050<br>(.045-.055) | M4<br>vs<br>M3 | 87.954<br>(25)<br>p=.000 | -.018 | .002 |

## Data Availability

The data can be found at figshare.com under the following DOI: https://doi.org/10.6084/m9.figshare.30236533

## Author Contributions

Conceptualization, H.C.P. and M.H.M.; methodology, H.C.P., M.H.M. and G.B.A.; software, M.H.M.; formal analysis, H.C.P. and M.H.M.; investigation, H.C.P., M.H.M.; G.B.A.; A.A.V. and S.H.F.; data curation, M.H.M., A.A.V. and G.B.A.; writing—original draft preparation, M.H.M., A.A.V. and S.H.F.; writing—review and editing, H.C.P., M.H.M.; G.B.A.; A.A.V. and S.H.F.; supervision, H.C.P.

